# Post-recovery health domain scores among outpatients by SARS-CoV-2 testing status during the pre-Delta period

**DOI:** 10.1101/2023.08.14.23294086

**Authors:** Jennifer P. King, Jessie R. Chung, James G. Donahue, Emily T. Martin, Aleda M. Leis, Arnold S. Monto, Manjusha Gaglani, Kayan Dunnigan, Chandni Raiyani, Sharon Saydah, Brendan Flannery, Edward A. Belongia

## Abstract

**Background:** Symptoms of COVID-19 including fatigue and dyspnea, may persist for weeks to months after SARS-CoV-2 infection. This study compared self-reported disability among SARS-CoV-2-positive and negative persons with mild to moderate COVID-19-like illness who presented for outpatient care before widespread COVID-19 vaccination.

**Methods:** Unvaccinated adults with COVID-19-like illness enrolled within 10 days of illness onset at three US Flu Vaccine Effectiveness Network sites were tested for SARS-CoV-2 by molecular assay. Enrollees completed an enrollment questionnaire and two follow-up surveys (7–24 days and 2–7 months after illness onset) online or by phone to assess illness characteristics and health status. The second follow-up survey included questions measuring global health, physical function, fatigue, and dyspnea. Scores in the four domains were compared by participants’ SARS-CoV-2 test results in univariate analysis and multivariable Gamma regression.

**Results:** During September 22, 2020 – February 13, 2021, 2,712 eligible adults were enrolled, 1,541 completed the first follow-up survey, and 650 completed the second follow-up survey. SARS-CoV-2-positive participants were more likely to report fever at acute illness but were otherwise comparable to SARS-CoV-2-negative participants. At first follow-up, SARS-CoV-2-positive participants were less likely to have reported fully or mostly recovered from their illness compared to SARS-CoV-2-negative participants. At second follow-up, no differences by SARS-CoV-2 test results were detected in the four domains in the multivariable model.

**Conclusion:** Self-reported disability was similar among outpatient SARS-CoV-2-positive and -negative adults 2–7 months after illness onset.

## Background

The clinical syndrome of COVID-19 is characterized by a dry cough, fever, and dyspnea.[1, 2] Additional symptoms including, headache, muscle aches, sore throat, and diarrhea are frequently reported. Intermediate and long-term effects of infection with SARS-CoV-2 are recognized as a spectrum of post-COVID-19 condition (PCC), also referred to as Long COVID [3–6]. Symptoms including fatigue and dyspnea may persist for weeks to months even in persons with mild-to-moderate acute illness.[7, 8] Evidence suggests that SARS-CoV-2 may cause long-lasting or permanent damage to the lungs and other organ systems following infection, as has been seen with SARS.[9, 10] The burden of PCC in the United States is substantial; by November 2021, among more than 46 million US adults estimated to have had COVID-19, 3–5 million experienced activity-limiting PCC.[11, 12] More recent surveys report that 11.2% of US adults who have ever had COVID-19 report PCC. [13]

This study employed four validated measures of global health, fatigue, physical function, and dyspnea to assess the level of self-reported disability 2–7 months after acute COVID-19 and compare disability between participants with COVID-19 and non-COVID-19 illness based on SARS-CoV-2 testing. The study was conducted before widespread COVID-19 vaccination.

## Methods

The source population for this study was adults (≥18 years of age) enrolled in a descriptive study of COVID-19 epidemiology at US Influenza Vaccine Effectiveness (Flu VE) network study sites in Michigan, Wisconsin, and Texas. We enrolled symptomatic persons seeking outpatient medical care (i.e., telehealth, primary care, urgent care, and emergency departments) or testing for SARS-CoV-2. All participants had COVID-19-like illness defined as acute respiratory illness that included fever, cough, or loss of taste or smell. Respiratory specimens collected within 10 days of illness onset were tested for SARS-CoV-2 using molecular assays [14]; results were used to classify participants as having COVID-19 or non-COVID-19 illness. COVID-19 vaccination status and presence of underlying health conditions prior to illness onset were extracted from the electronic medical record for all participants.

An enrollment questionnaire conducted in person or by phone collected pre-specified symptoms (shortness of breath/difficulty breathing, nasal congestion, chills, muscle aches, headache, vomiting, diarrhea, or abdominal pain), self-reported general health status, and demographic information. Two follow-up surveys were administered by email or over the phone. All participants were invited to complete the first follow-up survey approximately 7–24 days after illness onset [15]. The first survey included questions regarding self-reported symptoms, recovery date, if applicable, additional medical care required for the illness, and work productivity. Participants were aware of their SARS-CoV-2 status by the time of first follow-up survey initiation.

All participants who tested positive for SARS-CoV-2 at enrollment and a random sample of participants who tested negative were invited to complete the second follow-up survey approximately 2–7 months after illness onset. Roll out of the second follow-up survey varied by site and included participants enrolled from September 2020 through February 2021. To estimate the proportion of test-negative participants who became infected with SARS-CoV-2 before completion of the second follow-up survey, SARS-CoV-2 test-negative participants at one site were asked to provide a blood sample at the time of the second survey (28–42 days after illness onset) to test for anti-spike protein SARS-CoV-2 IgG antibodies (Beckman Coulter, Inc. Access SARS-CoV-2 IgG [16]).

The second follow-up survey assessed global health, fatigue, physical function and dyspnea using validated, standardized short form instruments from the Patient-Reported Outcomes Measurement Information System (PROMIS) network (https://www.healthmeasures.net/explore-measurement-systems/promis) (Supplemental Table 1).

The items in each instrument measure responses using a 5-point Likert-type scale. PROMIS short form instruments are scored using item-level calibrations and response pattern scoring which is considered more accurate than the use of raw scores. PROMIS uses a T-score– standardized metric in which 50 is the mean T-score of a relevant reference population (i.e., the US general population) and the standard deviation (SD) is 10. For PROMIS measures, higher scores represent a greater degree of the outcome being assessed (e.g., more fatigue). Formal studies of the various instruments in selected patient populations have determined that the short form instruments provided valid results. [17–23] This activity was reviewed and approved by US Centers for Disease Control and Prevention and each study site’s Institutional Review Board.

### Primary analysis

We stratified demographic characteristics, self-reported signs/symptoms at enrollment and other characteristics by SARS-CoV-2 test status. Participant characteristics were compared by χ^2^ tests, 2-sample t-tests, or Wilcoxon two-sample tests as appropriate. Standardized scores for global health, fatigue, physical function, and dyspnea were computed and compared by SARS-CoV-2 test results. Gamma regression was used to perform unadjusted and adjusted comparisons of mean T-scores as ratios (i.e., mean T-score among SARS-CoV-2 test-positive participants divided by mean T-score among SARS-CoV-2 test-negative participants) with corresponding 95% confidence intervals.[24] Separate models were fit for each PROMIS domain with the respective PROMIS T-score as the outcome variable. Participants who self-reported positive SARS-CoV-2 test results after enrollment were excluded. SARS-CoV-2 status (SARS-CoV-2 RT-PCR positive vs. negative at enrollment) was the main exposure variable for primary analyses. The base model was adjusted a priori for age (natural cubic spline with quintile knots), sex, interval between onset and follow-up survey (log-transformed days), and study site.

Additional potential confounders were evaluated using likelihood ratio tests. Adjustment for health-related factors included covariates for presence of any underlying chronic condition, self-reported cigarette smoking, body mass index, and self-rated general health status. Adjustment for socio-demographic factors included covariates for self-reported race and Hispanic ethnicity, and education level, along with receipt of 2020–21 seasonal influenza vaccine. An additional analysis was conducted on the subgroup of participants from one site for whom blood specimens during follow-up were collected; we excluded participants who had a negative RT-PCR result for SARS-CoV-2 at enrollment but who tested SARS-CoV-2 seropositive 28–42 days after the enrollment illness onset.

Participants were excluded if they did not complete all PROMIS instruments, had uninterpretable SARS-CoV-2 test results at enrollment or if they received ≥1 dose of any COVID-19 vaccine before the enrollment illness onset (Supplemental Figure 1).

### Sensitivity Analyses

We conducted three additional analyses to assess the robustness of our primary findings. First, we restricted the analysis to participants who reported fever at enrollment. Second, we restricted the analysis to participants with underlying conditions in three or more categories. Third, we excluded participants who reported they had fully or mostly recovered at the short-term follow-up assessment.

### Sample size

A total of 200 SARS-CoV-2 test-positive and 200 SARS-CoV-2 test-negative participants, would allow detection of an effect size (differences in mean T-scores) of 2.8 with 80% power. This corresponds to a standardized effect size ([group_1_ mean – group_2_ mean]/[SD]) of 0.28 which is generally considered small to moderate in size. Other parameters in the calculation were α=0.05 and SD of the outcome in the population=10.

## Results

Between September 22, 2020 and February 13, 2021, 2,712 adults were enrolled at the three sites. Of those, 1,541 (57%) completed the first follow-up survey. Of 1,350 participants invited to complete the second survey, 650 (48%) attempted it, and 578 (89%) were included in this analysis, including 312 participants in the SARS-CoV-2 test-positive group and 266 participants in the SARS-CoV-2 test-negative group. None of the participants tested positive for influenza at enrollment. Participants were enrolled a median of 9 days after illness onset (interquartile range (IQR), 6–14). SARS-CoV-2 test-positive participants reported a mean of 4.5 (SD, 1.6) signs/symptoms at enrollment compared to a mean of 3.8 (SD, 1.6) among SARS-CoV-2 test-negative participants (Table 1). We observed differences in the signs/symptoms reported by participants by SARS-CoV-2 test result. Of note, a greater proportion of SARS-CoV-2-positive participants reported loss of sense of taste/smell compared to SARS-CoV-2 test-negative participants (65% vs 21%, p<0.01). Among other differences, a greater proportion (74%) of participants in the SARS-CoV-2 test-positive group reported fever or chills compared to participants in the SARS-CoV-2 test-negative group (65%) (p=0.02).

**Table 1.** Illness characteristics among participants in the SARS-CoV-2-positive and SARS-CoV-2-negative groups, N (%)

|  | Positive SARS-CoV-2 result |  | Negative SARS-CoV-2 result |  | p-value |
| --- | --- | --- | --- | --- | --- |
|  | N | % | N | % |  |
| **N** | 312 | 100 | 266 | 100 |  |
| **Total number of symptoms, mean (SD)** | 4.5 | 1.6 | 3.8 | 1.6 | <0.01 |
| **Reported symptoms at enrollment** |  |  |  |  |  |
| *Any respiratory symptom* | 309 | 99 | 242 | 91 | <0.01 |
| Cough | 258 | 83 | 201 | 76 | 0.03 |
| Loss of taste or smell | 202 | 65 | 57 | 21 | <0.01 |
| Shortness of breath | 124 | 40 | 82 | 31 | 0.02 |
| Congestion/runny nose<sup>1</sup> | 212 | 86 | 164 | 74 | <0.01 |
| Sore throat | 157 | 50 | 158 | 59 | 0.03 |
| *Any generalized sign or symptom* | 300 | 96 | 244 | 92 | 0.01 |
| Fever or chills | 231 | 74 | 173 | 65 | 0.02 |
| Fatigue | 209 | 67 | 178 | 67 | 0.21 |
| Muscle aches | 220 | 71 | 140 | 53 | <0.01 |
| Headache | 247 | 79 | 182 | 68 | <0.01 |
| *Any gastrointestinal symptom* | 166 | 53 | 126 | 47 | 0.15 |
| Nausea/vomiting | 84 | 27 | 87 | 33 | 0.13 |
| Diarrhea | 136 | 44 | 82 | 31 | <0.01 |
| **Sought subsequent medical care for illness<sup>2</sup>** | 16 | 6 | 18 | 8 | 0.51 |
| **Reported recovery at 1st follow up survey<sup>3</sup>** | 197 | 80 | 197 | 90 | <0.01 |

SD, standard deviation
<sup>1</sup> Not assessed in 111 participants including 66 SARS-CoV-2 test-positive participants and 164 SARS-CoV-2 test-negative participants.<sup>2</sup> Whether the participant sought medical care for their illness after enrollment as reported at the first follow-up survey. Missing for 41 SARS-CoV-2 test-negative participants and 63 SARS-CoV-2 test-positive participants.<sup>3</sup> Participants who responded “yes” to the question “Have you fully or mostly recovered from your illness?” on the first follow-up survey. Missing for 46 SARS-CoV-2 test-negative participants and 66 SARS-CoV-2 test-positive participants.

The median time from illness onset to first follow-up survey was 21 days (IQR 14–30). Participants in the SARS-CoV-2 test-positive group had a shorter interval between onset and first follow-up survey completion (median 16.5 days, IQR 14–26) than participants in the SARS-CoV-2 test-negative group (median 26 days, IQR 16–33) (p<0.01). On the first follow-up survey, 80% of participants in the SARS-CoV-2 test-positive group reported they had mostly or fully recovered from their illness compared to 90% of participants in the SARS-CoV-2 test-negative group. The median reported time between illness onset and date of recovery was 12 days (IQR 10–17) among participants in the SARS-CoV-2 test-positive group compared to 9 days (IQR 5– 13) among participants in the SARS-CoV-2 test-negative group. Among participants who reported they had not yet recovered from the acute illness on the first follow-up survey, 19 (73%) participants who tested SARS-CoV-2 positive reported on-going fatigue compared to 7 (54%) participants who tested SARS-CoV-2 negative (p=0.23). Reporting of fever or chills was the same (15%) for both groups.

Among participants who completed the second follow-up survey, mean age was 47.2 years (SD 14.7); participants in the SARS-CoV-2 test-negative group were more likely to be female, a self-reported smoker, vaccinated against influenza, have a higher education level, and were slightly younger (mean age 45.7 years, SD 14.7) than participants in the SARS-CoV-2 test-positive group (mean age 48.5 years, SD 14.6) (p=0.02) (Table 2). Participants completed the second follow-up survey approximately three months (median 89 days, IQR 72–111) after illness onset. Participants in the SARS-CoV-2 test-positive group had a shorter interval between onset and second follow-up survey completion (median 89 days, IQR 74–119) than participants in the SARS-CoV-2 test-negative group (median 104 days, IQR 71–125) (p<0.01).

**Table 2.** Demographic and other characteristics of SARS-CoV-2 test-positive and SARS-CoV-2 test-negative participants who completed the second follow-up survey.

|  | Total |  | Positive SARS-CoV-2 result |  | Negative SARS-CoV-2 result |  | P-value<sup>1</sup> |
| --- | --- | --- | --- | --- | --- | --- | --- |
|  | N<sup>2</sup> | %<sup>2</sup> | N<sup>2</sup> | %<sup>2</sup> | N<sup>2</sup> | %<sup>2</sup> |  |
| **N** | 578 | 100 | 312 | 100 | 266 | 100 |  |
| **Study site** |  |  |  |  |  |  | <0.01 |
| Michigan | 277 | 48 | 107 | 34 | 170 | 64 |  |
| Texas | 111 | 19 | 66 | 21 | 45 | 17 |  |
| Wisconsin | 190 | 33 | 139 | 45 | 51 | 19 |  |
| **Female sex<sup>3</sup>** | 398 | 69 | 204 | 65 | 194 | 73 | 0.04 |
| **Age (years)** |  |  |  |  |  |  | 0.19 |
| Mean (SD) | 47.2 | 14.7 | 48.5 | 14.6 | 45.7 | 14.7 |  |
| 18–49 | 303 | 52 | 153 | 49 | 150 | 56 |  |
| 50–64 | 203 | 35 | 119 | 38 | 84 | 32 |  |
| ≥65 | 72 | 12 | 40 | 13 | 32 | 12 |  |
| **Race/ethnicity<sup>4</sup>** |  |  |  |  |  |  | 0.78 |
| White, non-Hispanic | 505 | 87 | 274 | 88 | 231 | 87 |  |
| Black, non-Hispanic | 10 | 2 | 5 | 2 | 5 | 2 |  |
| Other, non-Hispanic | 28 | 5 | 13 | 4 | 15 | 6 |  |
| Hispanic, any race | 32 | 6 | 19 | 6 | 13 | 5 |  |
| **Body Mass Index<sup>5</sup>** |  |  |  |  |  |  | 0.24 |
| Median (IQR) | 29.7 | 25.6–35.1 | 30.3 | 25.9–35.4 | 29.4 | 24.8–34.7 |  |
| Underweight (<18.5) | 2 | <1 | 1 | 0.3 | 1 | <1 |  |
| Normal (18.5–24) | 115 | 20 | 52 | 17 | 63 | 24 |  |
| Overweight (25–29) | 165 | 29 | 91 | 29 | 74 | 28 |  |
| Obese (30–39) | 201 | 35 | 118 | 38 | 83 | 31 |  |
| Morbidly Obese (≥40) | 73 | 13 | 39 | 13 | 34 | 13 |  |
| **Self-reported underlying health condition<sup>6</sup>** | 204 | 35 | 107 | 34 | 97 | 36 | 0.55 |
| **Documented underlying health condition** |  |  |  |  |  |  |  |
| ≥1 condition | 340 | 59 | 187 | 60 | 153 | 58 | 0.58 |
| ≥3 conditions | 123 | 21 | 62 | 20 | 61 | 23 | 0.50 |
| Metabolic disease | 156 | 27 | 82 | 26 | 74 | 28 |  |
| Hypertension | 131 | 23 | 73 | 23 | 58 | 22 |  |
| Chronic pulmonary disease | 78 | 13 | 40 | 13 | 38 | 14 |  |
| Neurological/musculoskeletal | 64 | 11 | 27 | 9 | 37 | 14 |  |
| Endocrine disorder | 63 | 11 | 38 | 12 | 25 | 9 |  |
| Chronic cardiac disease | 61 | 11 | 32 | 10 | 29 | 11 |  |
| Diabetes mellitus | 61 | 11 | 38 | 12 | 23 | 9 |  |
| Malignancy | 51 | 9 | 25 | 8 | 26 | 10 |  |
| Chronic renal disease | 31 | 5 | 18 | 6 | 13 | 5 |  |
| Immunosuppressive disorder | 30 | 5 | 15 | 5 | 15 | 6 |  |
| Other condition<sup>7</sup> | 24 | 4 | 13 | 4 | 11 | 4 |  |
| Liver disease | 23 | 4 | 12 | 4 | 11 | 4 |  |
| **Self-rated general health status** |  |  |  |  |  |  | <0.01 |
| Excellent | 139 | 24 | 73 | 23 | 66 | 25 |  |
| Very Good or Good | 413 | 71 | 233 | 75 | 180 | 68 |  |
| Fair or Poor | 26 | 5 | 6 | 2 | 20 | 8 |  |
| **Received 2020–21 influenza vaccine<sup>8</sup>** | 375 | 65 | 190 | 61 | 185 | 70 | 0.02 |
| **Cigarette smoking** |  |  |  |  |  |  | 0.04 |
| Every day or some days | 39 | 7 | 15 | 5 | 24 | 9 |  |
| Not at all | 539 | 93 | 297 | 95 | 242 | 91 |  |
| **Education** |  |  |  |  |  |  | <0.01 |
| Less than high school/high school graduate/GED | 77 | 13 | 52 | 17 | 25 | 10 |  |
| Some college<sup>9</sup> | 177 | 31 | 106 | 34 | 71 | 27 |  |
| Bachelor's degree | 191 | 33 | 94 | 30 | 97 | 36 |  |
| Advanced degree | 133 | 23 | 60 | 19 | 73 | 27 |  |
| **Month of illness onset** |  |  |  |  |  |  | <0.01 |
| September 2020 | 38 | 7 | 4 | 1 | 34 | 13 |  |
| October 2020 | 110 | 19 | 42 | 13 | 68 | 26 |  |
| November 2020 | 298 | 52 | 198 | 63 | 100 | 38 |  |
| December 2020 | 52 | 9 | 28 | 9 | 24 | 9 |  |
| January 2021 | 77 | 13 | 39 | 13 | 38 | 14 |  |
| February 2021 | 3 | 1 | 1 | 0.9 | 2 | 1 |  |
| **Interval between onset and 2nd follow up, median days (IQR)** | 89 | 72–111 | 89 | 74–119 | 104 | 71–125 | <0.01 |

GED, general education degree; IQR, interquartile range; SD, standard deviation
<sup>1</sup> P-value for the comparison between SARS-CoV-2 test-positive and test negative participants
<sup>2</sup> Cell numbers are number or column percent unless otherwise specified.
<sup>3</sup> Missing for 1 SARS-CoV-2 test-negative participant
<sup>4</sup> Self-reported at enrollment. Other race includes participants who selected Asian, Native Hawaiian/Other Pacific Islander, American Indian/Alaska Native, or Other and participants who selected more than one race. Missing for 2 SARS-CoV-2 test-negative and 1 SARS-CoV-2 test-positive participants.
<sup>5</sup> Missing for 11 SARS-CoV-2 test-negative and 11 SARS-CoV-2 test-positive participants.
<sup>6</sup> Self-reported at enrollment to have any of the following: heart disease, lung disease, diabetes, cancer, liver or kidney disease, immune suppression, or high blood pressure. Missing for 4 SARS-CoV-2 test-negative and 2 SARS-CoV-2 test-positive participants.
<sup>7</sup> Includes hemoglobinopathies, cerebrovascular disease, and disease of arteries, arterioles, and capillaries.
<sup>8</sup> Seasonal influenza vaccination receipt as self-reported at enrollment. Missing for 2 SARS-CoV-2 test-negative and 1 SARS-CoV-2 test-positive participants.
<sup>9</sup> Includes vocational training or associate degree

At the second follow-up time point, there was no overall difference between the groups in unadjusted mean T-scores in the domains of global health, physical function, or dyspnea; however, participants in the SARS-CoV-2 test-negative group reported more fatigue than participants in the SARS-CoV-2 test-positive group (Figure 1, Supplemental Table 2). Among participants in the SARS-CoV-2 test-negative group, unadjusted mean T-scores (SD) for the global health, physical function, fatigue, and dyspnea domains were 51.7 (7.4), 52.2 (7.1), 46.9 (10.2), and 40.6 (7.9), respectively. Among participants in the SARS-CoV-2 test-positive group, mean T-scores (SD) for the global health, physical function, fatigue, and dyspnea domains were 51.9 (7.2), 52.1 (7.2), 44.6 (9.4), and 40.1 (7.1), respectively.

**Figure 1.**
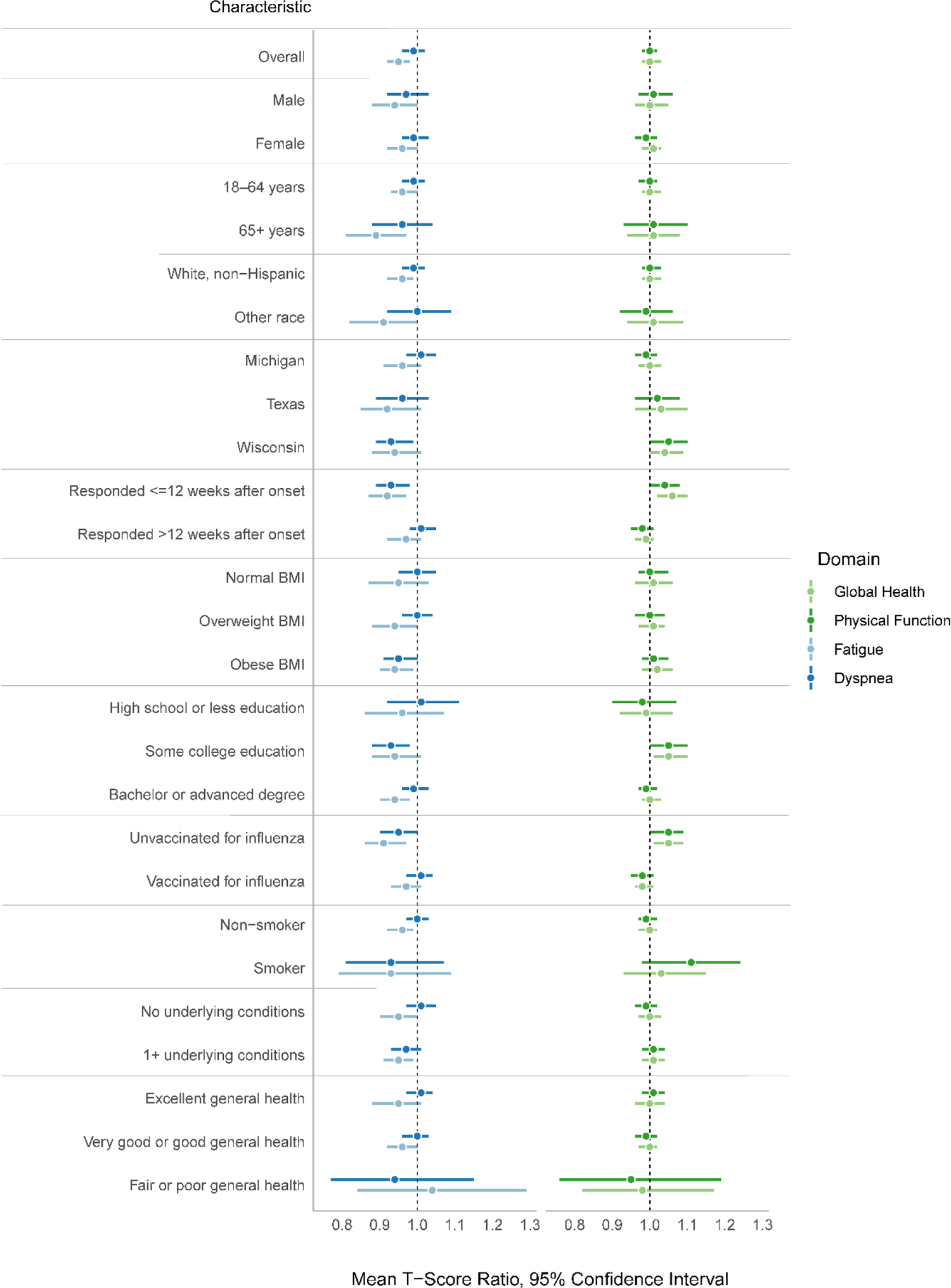
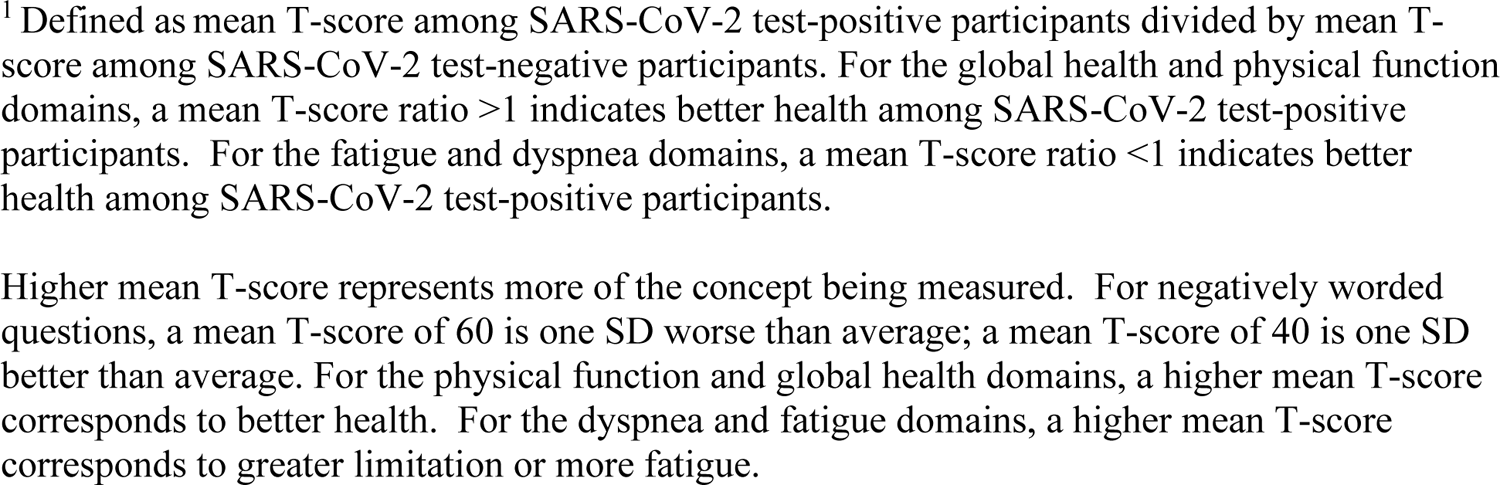
Unadjusted Global Health, Physical Function, Fatigue, and Dyspnea domain mean T-score ratios^1^ stratified by participant characteristics

We observed statistically significant mean T-score ratios within subgroups of participants (e.g., among those who completed the second follow-up survey within three months after illness onset), which was generally in the same direction within a domain. Among participants who completed the second follow-up survey within three months of illness onset, participants in the SARS-CoV-2 test-negative group reported more impairment on their health and function compared to participants in the SARS-CoV-2 test-positive group. Differences were not observed among participants who completed the second follow-up survey more than 3 months after illness onset.

After adjustment for participant age at enrollment, participant sex, interval between onset and 2^nd^ follow-up survey completion, study site, presence of any underlying health condition, participant-reported cigarette smoking, and self-rated general health status, we observed no statistically significant mean T-score ratios in any domain (Figure 2). The addition of social factors to the model including base and health factors did not improve model fit or change interpretation of findings (Supplemental Figure 2).

**Figure 2.**
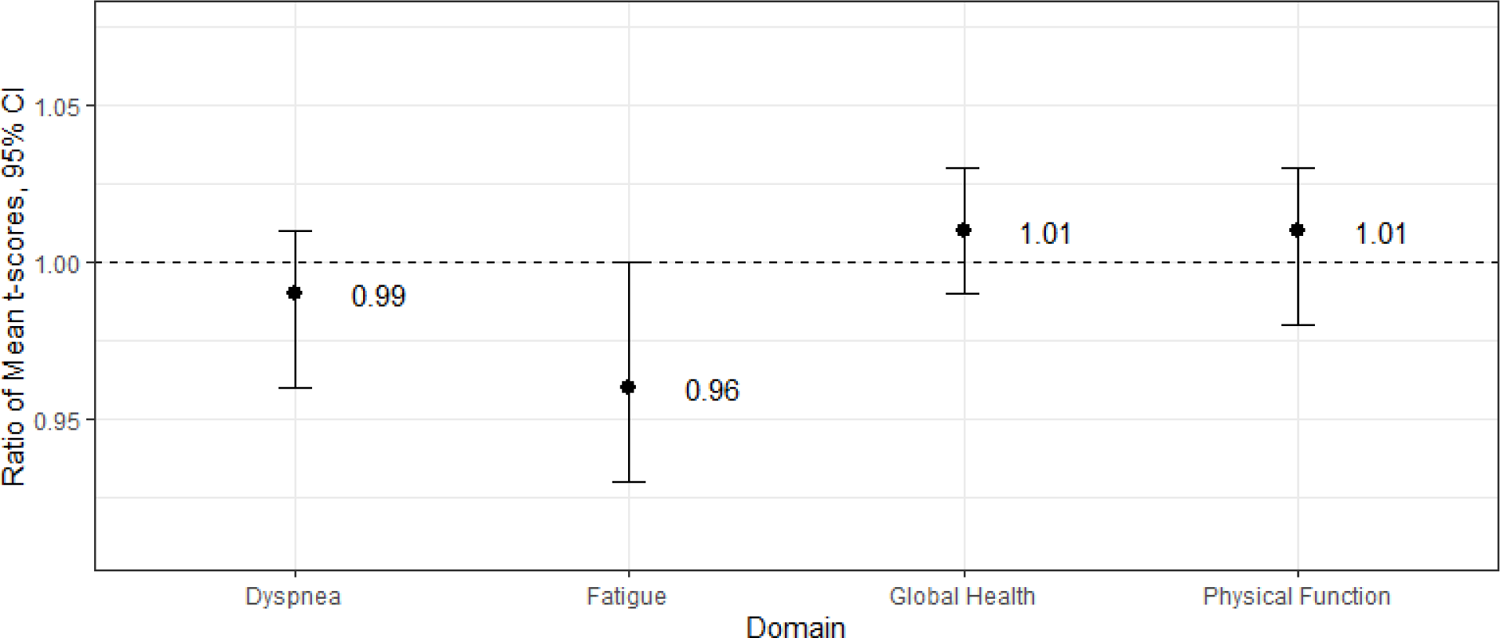
Adjusted Global Health, Physical Function, Fatigue, and Dyspnea domain mean T-score ratios from multivariable^1^ Gamma regression.

Among 51 SARS-CoV-2 test-negative participants from the Wisconsin site who had SARS-CoV-2 serologic testing, 5 (10%) tested seropositive at second follow-up that occurred during the same pre-Delta variant period. These participants were similar to other participants in the SARS-CoV-2 test-negative group with respect to age, sex, and month of illness onset; reported symptoms at enrollment among the five SARS-CoV-2 seropositive participants included cough (n=5), fever (n=3), and sore throat (n=2). Adjusted mean T-score ratios were similar for all domains, when the five participants who were seropositive in the SARS-CoV-2 test-negative group were excluded (Supplemental Table 3).

Adjusted mean T-score ratios in the three sensitivity analyses were generally consistent with primary analyses (Supplemental Table 3). In the sensitivity analysis restricted to participants with underlying conditions in three or more categories, SARS-CoV-2 test-positive participants tended to experience worse outcomes for physical function and fatigue but these differences were not statistically significant.

## Discussion

In this observational study of adults seeking outpatient medical care for an acute symptomatic respiratory illness from September 2020 through February 2021 when COVID-19 vaccines were not yet widely available, there was no difference between SARS-CoV-2 test-positive compared to SARS-CoV-2 test-negative participants surveyed 2–7 months after illness onset in self-reported global health, fatigue, physical function, or dyspnea as measured using four validated PROMIS domains. These findings contribute to evidence that prevalence of symptoms and conditions consistent with PCC may not be limited to post-SARS-CoV-2 respiratory viral infection. This study had several strengths including systematic testing to confirm SARS-CoV-2 status, a geographically diverse study population, and the use of standardized instruments.

The prevalence of persistent symptoms more than two months after illness onset and overall effects on well-being following acute illness found in this study are within the range reported from other studies following patients diagnosed with medically attended COVID-19 or mildly symptomatic SARS-CoV-2 infection. [25–27] Three similar studies, including one that also measured specific health domains, reported comparable results when SARS-CoV-2 test-positive patients were compared to SARS-CoV-2 test-negative patients. [28–30] While acute symptoms and quality of life indicators may differ between ambulatory patients with and without SARS-CoV-2, there is substantial overlap in the clinical features of infection caused by SARS-CoV-2 and other respiratory viruses, such as influenza. [14, 31] Participants in both groups in this analysis were more likely to report being fully or mostly recovered from their illness on the first follow-up survey compared to what was reported among adults enrolled in the US Flu Vaccine Effectiveness Network during the 2017–18 influenza season. In that pre-COVID-19 pandemic influenza season when approximately one-third of participants tested positive for influenza, 32% of adults aged 19–64 years who completed the follow-up survey reported they had not yet fully or mostly recovered 7–21 days after illness onset. [32] In that influenza season, the median time between illness onset and recovery among participants who had fully or mostly recovered at follow-up was 11 days, similar to median duration of illness observed in this study (9 days for SARS-CoV-2 test-negative and 12 days for SARS-CoV-2 test-positive participants).

It remains unclear how prevalent long-term sequelae are with respect to other common respiratory viral pathogens and why SARS-CoV-2 infection can result in long-term sequelae in some individuals but not in others. Introduction of zoonotic coronavirus infection in naïve human populations with limited cross-protection from common human coronaviruses may have increased pathogenicity or intensity of human immune response until SARS-CoV-2 adapted to human hosts and the population developed partial immunity. Alternatively, the magnitude of COVID-19 cases may have increased attention to post-viral syndromes and persistence of symptoms common to many viral infections, including fatigue and persistent decrease in lung function. Early recognition of persistent or new symptoms, including fatigue, months after laboratory-confirmed COVID-19 may have increased awareness of and healthcare seeking for PCC.[33] Many early reports and studies described more severe post-COVID-19 syndromes following severe and prolonged acute illness. [34, 35] The description of and evidence for less severe PCC following even mild symptomatic COVID-19 followed from cohort studies and suggested that most mild illness was self-limited. Findings were similar in the sensitivity analysis in this study that was restricted to outpatients who reported fever. Because persistent symptoms occur following many viral infections and infections may exacerbate underlying chronic conditions with similar clinical presentation, inclusion of a comparison group of patients with mild illness who test negative for acute SARS-CoV-2 infection is needed to identify specific characteristics of post-SARS-CoV-2 infection sequelae.

The findings presented here are subject to several limitations. First, our findings may not be generalizable to the wider population of adults who seek care for mild to moderate COVID-19. During the period of this study, there could have been differences in the people who were seeking care due to restrictions on in-person medical encounters. Persons who presented for care during this time might have had more underlying medical conditions or other unmeasured differences. Further, many patients approached for participation in the follow-up surveys following initial SARS-CoV-2 test declined, leading to a non-representative and potentially biased sample of all symptomatic patients. Second, we did not test controls for other etiologies besides SARS-CoV-2 and influenza viruses. Finally, our comparison group may have been previously infected with SARS-CoV-2 or infected before the follow-up surveys after testing negative for infection at acute illness. Serology conducted only at one site indicated that approximately 10% of SARS-CoV-2 test-negative patients enrolled at that site had been infected before the second follow-up survey. While excluding these patients did not change results, inclusion of patients with SARS-CoV-2 in the comparison group would bias results towards the null.

These results highlight that many individuals may continue to experience on-going symptoms in the weeks following an acute respiratory infection and that these symptoms are likely not unique to SARS-CoV-2 infection. Characterization of PCC remains challenging as immune and vaccine history grows more complex and with the emergence of new SARS-CoV-2 variants. Our study was conducted during a unique time during the COVID-19 pandemic.

Importantly, the study period preceded widespread COVID-19 vaccination and emergence of the Delta and Omicron variants when patients may have had multiple SARS-CoV-2 infections. As a result, the unvaccinated participants who tested SARS-CoV-2 positive at enrollment were likely infected for the first time. Widespread COVID-19 vaccination efforts and booster campaigns may reduce the occurrence of PCC or modify its characteristics.[36] Inclusion of comparison groups of symptomatic patients with medically attended illness in future evaluations of PCC will help identify contributing SARS-CoV-2-specific factors versus non-specific factors that could be targeted with different interventions.

## Data Availability

All data produced in the present study are available upon reasonable request to the authors.

---

**Supplemental Figure 1.**
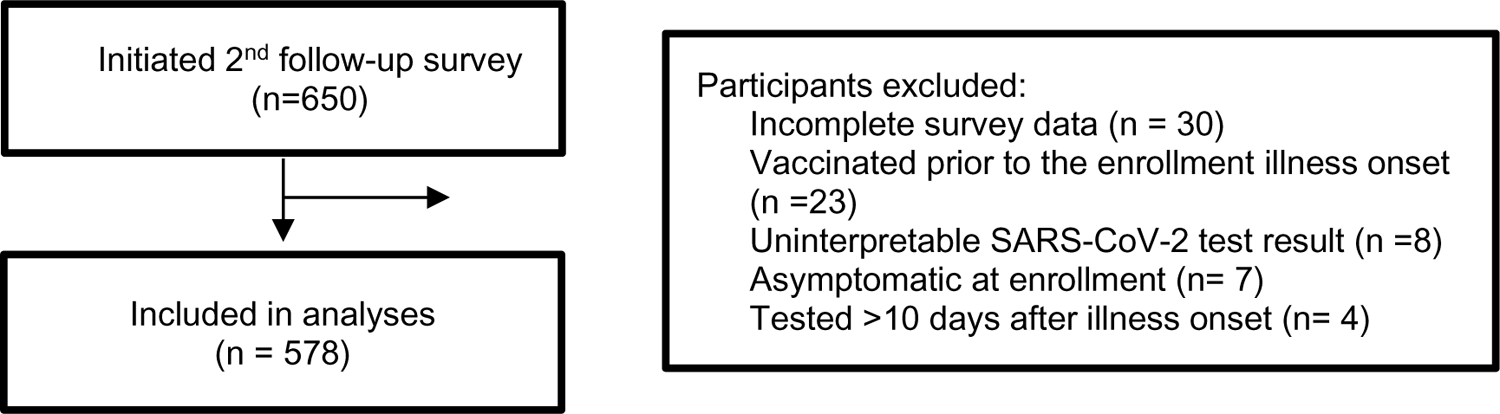

**Supplemental Figure 2.**
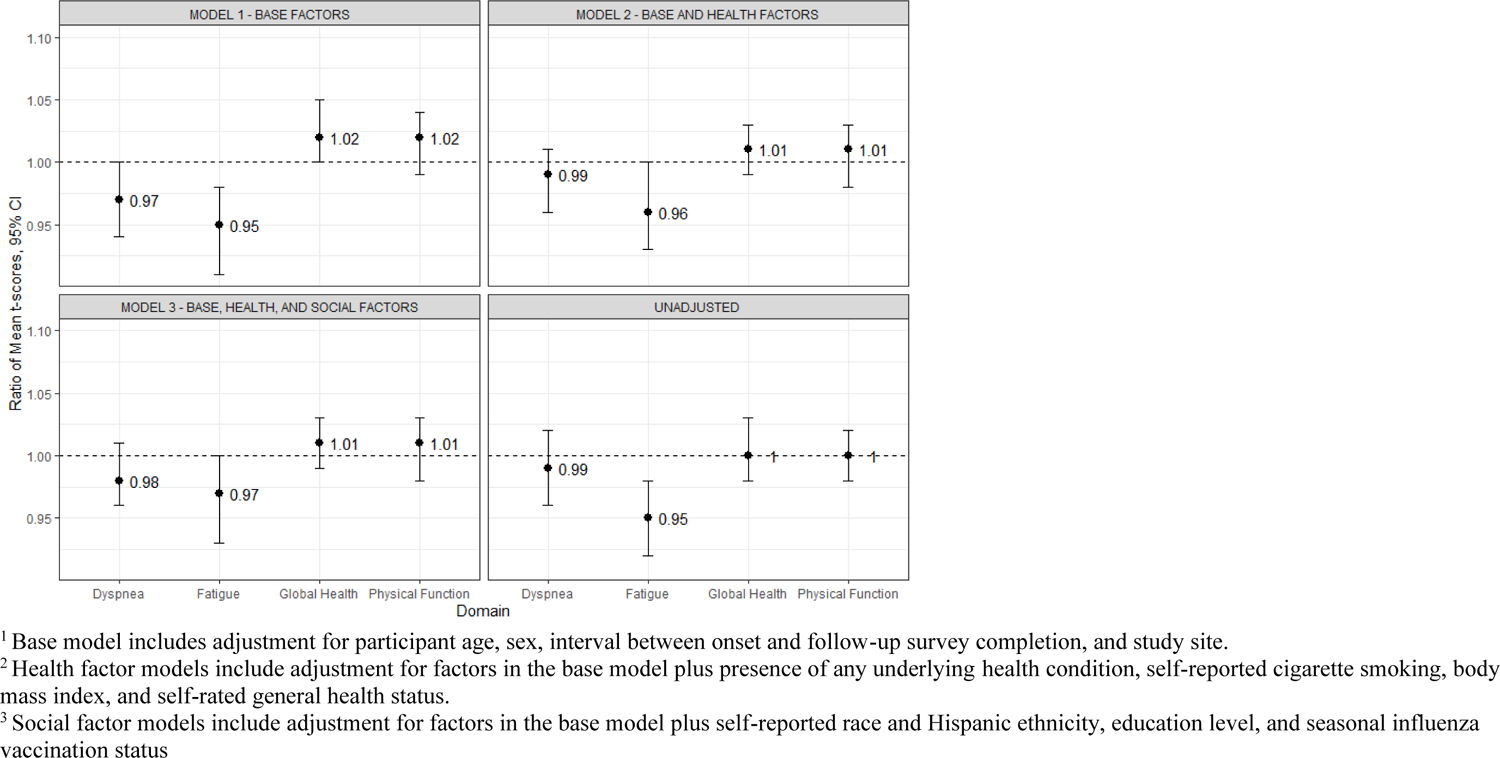
Adjusted Global Health, Physical Function, Fatigue, and Dyspnea domain mean T-score ratios from multivariable^1-3^ Gamma regression models.

**Supplemental Table 1.** PROMIS questions administered on second follow-up survey.

| Domain | Question | Answer choices |
| --- | --- | --- |
| Global Health | In general, how would you rate your physical health? | 1 – Poor<br>2 – Fair<br>3 – Good<br>4 – Very good<br>5 – Excellent |
|  | To what extent are you able to carry out your everyday physical activities? | 1 – Not at all<br>2 – A little<br>3 – Moderately<br>4 – Mostly<br>5 – Completely |
| Physical Function | Are you able to do chores such as vacuuming or yard work? | 1 – Unable to do<br>2 – With much difficulty<br>3 – With some difficulty<br>4 – With a little difficulty<br>5 – Without any difficulty |
|  | Are you able to go up and down stairs at a normal pace? | 1 – Unable to do<br>2 – With much difficulty<br>3 – With some difficulty<br>4 – With a little difficulty<br>5 – Without any difficulty |
|  | Are you able to go for a walk of at least 15 minutes? | 1 – Unable to do<br>2 – With much difficulty<br>3 – With some difficulty<br>4 – With a little difficulty<br>5 – Without any difficulty |
|  | Are you able to run errands and shop? | 1 – Unable to do<br>2 – With much difficulty<br>3 – With some difficulty<br>4 – With a little difficulty<br>5 – Without any difficulty |
| Dyspnea Functional Limitation | Considering your shortness of breath over the past 7 days, rate the amount of difficulty you had when doing the following activities: |  |
|  | Dressing yourself without help | 0 – No difficulty<br>1 – A little difficulty<br>2 – Some difficulty<br>3 – Much difficulty<br>NA – I did not do this in the past 7 days |
|  | Walking 50 steps/paces on flat ground at a normal speed without stopping | 0 – No difficulty<br>1 – A little difficulty<br>2 – Some difficulty<br>3 – Much difficulty<br>NA – I did not do this in the past 7 days |
|  | Walking up 20 stairs (2 flights) without stopping | 0 – No difficulty<br>1 – A little difficulty<br>2 – Some difficulty<br>3 – Much difficulty<br>NA – I did not do this in the past 7 days |
|  | Preparing meals | 0 – No difficulty<br>1 – A little difficulty<br>2 – Some difficulty<br>3 – Much difficulty |
|  |  | NA – I did not do this in the past 7 days |
|  | Washing dishes | 0 – No difficulty<br>1 – A little difficulty<br>2 – Some difficulty<br>3 – Much difficulty<br>NA – I did not do this in the past 7 days |
|  | Sweeping or mopping | 0 – No difficulty<br>1 – A little difficulty<br>2 – Some difficulty<br>3 – Much difficulty<br>NA – I did not do this in the past 7 days |
|  | Making a bed | 0 – No difficulty<br>1 – A little difficulty<br>2 – Some difficulty<br>3 – Much difficulty<br>NA – I did not do this in the past 7 days |
|  | Lifting something weighing 10-20 lbs<br>(about 4.5-9 kg, like a large bag of groceries) | 0 – No difficulty<br>1 – A little difficulty<br>2 – Some difficulty<br>3 – Much difficulty<br>NA – I did not do this in the past 7 days |
|  | Carrying something weighing 10-20 lbs<br>(about 4.5-9 kg, like a large bag of groceries) from one room to another | 0 – No difficulty<br>1 – A little difficulty<br>2 – Some difficulty<br>3 – Much difficulty<br>NA – I did not do this in the past 7 days |
|  | Walking (faster than your usual speed)<br>for ½ mile (almost 1 km) without<br>stopping | 0 – No difficulty<br>1 – A little difficulty<br>2 – Some difficulty<br>3 – Much difficulty<br>NA – I did not do this in the past 7 days |
| Fatigue | During the past 7 days:<br>I feel fatigued | 1 – Not at all<br>2 – A little bit<br>3 – Somewhat<br>4 – Quite a bit<br>5 – Very much |
|  | I have trouble <u>starting</u> things because I<br>am tired | 1 – Not at all<br>2 – A little bit<br>3 – Somewhat<br>4 – Quite a bit<br>5 – Very much |
|  | In the past 7 days: |  |
|  | How run-down did you feel on average? | 1 – Not at all<br>2 – A little bit<br>3 – Somewhat<br>4 – Quite a bit<br>5 – Very much |
|  | How fatigued were you on average? | 1 – Not at all<br>2 – A little bit<br>3 – Somewhat<br>4 – Quite a bit<br>5 – Very much |

NA, not applicable

**Supplemental Table 2.** Unadjusted raw PROMIS mean T-scores^1^ and mean T-score ratios^2^ by SARS-CoV-2 test-positive or SARS-CoV-2 test-negative groups and participant characteristics.

|  | Global health<sup>3</sup> |  |  | Physical function<sup>3</sup> |  |  | Fatigue<sup>4</sup> |  |  | Dyspnea<sup>4</sup> |  |  |
| --- | --- | --- | --- | --- | --- | --- | --- | --- | --- | --- | --- | --- |
|  | SARS-CoV-2 negative | SARS-CoV-2 positive | Ratio (95% CI) | SARS-CoV-2 negative | SARS-CoV-2 positive | Ratio (95% CI) | SARS-CoV-2 negative | SARS-CoV-2 positive | Ratio (95% CI) | SARS-CoV-2 negative | SARS-CoV-2 positive | Ratio (95% CI) |
| **Overall** | 51.7<br>(7.4) | 51.9<br>(7.2) | 1.00<br>(0.98-1.03) | 52.2<br>(7.1) | 52.1<br>(7.2) | 1.00<br>(0.98-1.02) | **46.9**<br>**(10.2)** | **44.6**<br>**(9.4)** | **0.95**<br>**(0.92-0.98)** | 40.6<br>(7.9) | 40.1<br>(7.1) | 0.99<br>(0.96-1.02) |
| **Sex** |  |  |  |  |  |  |  |  |  |  |  |  |
| Male | 51.2<br>(8.0) | 51.4<br>(7.1) | 1.00<br>(0.96-1.05) | 52.2<br>(7.3) | 52.9<br>(7.4) | 1.01<br>(0.97-1.06) | 46.4<br>(9.6) | 43.6<br>(9.4) | 0.94<br>(0.88-1.00) | 40.6<br>(8.3) | 39.5<br>(6.9) | 0.97<br>(0.92-1.03) |
| Female | 51.8<br>(7.2) | 52.1<br>(7.2) | 1.01<br>(0.98-1.03) | 52.1<br>(7.0) | 51.7<br>(7.1) | 0.99<br>(0.96-1.02) | 47.2<br>(10.4) | 45.1<br>(9.4) | 0.96<br>(0.92-1.00) | 40.6<br>(7.7) | 40.3<br>(7.2) | 0.99<br>(0.96-1.03) |
| **Age group (years)** |  |  |  |  |  |  |  |  |  |  |  |  |
| 18–64 | 51.8<br>(7.4) | 52.0<br>(7.2) | 1.00<br>(0.98-1.03) | 52.6<br>(6.8) | 52.6<br>(6.9) | 1.00<br>(0.97-1.02) | 46.8<br>(10.4) | 45.0<br>(9.4) | 0.96<br>(0.93-1.00) | 40.1<br>(7.7) | 39.8<br>(7.0) | 0.99<br>(0.96-1.02) |
| 65+ | 50.5<br>(7.1) | 51.0<br>(7.3) | 1.01<br>(0.94-1.08) | 49.0<br>(8.1) | 49.4<br>(9.0) | 1.01<br>(0.93-1.10) | **47.6**<br>**(8.3)** | **42.2**<br>**(9.7)** | **0.89**<br>**(0.81-0.97)** | 43.8<br>(8.4) | 41.9<br>(7.5) | 0.96<br>(0.88-1.04) |
| **Race/ethnicity** |  |  |  |  |  |  |  |  |  |  |  |  |
| White, non-Hispanic | 52.0<br>(7.2) | 52.2<br>(7.1) | 1.00<br>(0.98-1.03) | 52.3<br>(7.0) | 52.3<br>(7.2) | 1.00<br>(0.98-1.03) | **46.3**<br>**(10.0)** | **44.4**<br>**(9.5)** | **0.96**<br>**(0.92-0.99)** | 40.6<br>(7.9) | 40.0<br>(7.0) | 0.99<br>(0.96 - 1.02) |
| Other | 49.1<br>(8.2) | 49.7<br>(7.6) | 1.01<br>(0.94-1.09) | 51.6<br>(7.6) | 50.9<br>(7.6) | 0.99<br>(0.92-1.06) | 50.7<br>(10.9) | 46.0<br>(9.2) | 0.91<br>(0.82-1.00) | 40.6<br>(7.4) | 40.7<br>(7.8) | 1.00<br>(0.92-1.09) |
| **Site** |  |  |  |  |  |  |  |  |  |  |  |  |
| Michigan | 53.3<br>(6.5) | 53.3<br>(6.2) | 1.00<br>(0.97-1.03) | 53.5<br>(6.3) | 53.0<br>(6.4) | 0.99<br>(0.96-1.02) | 46.2<br>(9.6) | 44.3<br>(9.2) | 0.96<br>(0.91-1.01) | 39.2<br>(7.0) | 39.4<br>(6.7) | 1.01<br>(0.97-1.05) |
| Texas | 46.7<br>(7.8) | 48.0<br>(8.3) | 1.03<br>(0.96-1.10) | 49.9<br>(8.2) | 50.7<br>(7.6) | 1.02<br>(0.96-1.08) | 49.4<br>(12.0) | 45.5<br>(10.0) | 0.92<br>(0.85-1.01) | 43.4<br>(10.0) | 41.5<br>(8.0) | 0.96<br>(0.89-1.03) |
| Wisconsin | **50.5**<br>**(7.6)** | **52.6**<br>**(6.7)** | **1.04**<br>**(1.00-1.09)** | **49.8**<br>**(7.5)** | **52.2**<br>**(7.6)** | **1.05**<br>**(1.00-1.10)** | 47.0<br>(10.2) | 44.4<br>(9.4) | 0.94 (0.88-1.01) | **42.7**<br>**(7.3)** | **39.9**<br>**(7.0)** | **0.93**<br>**(0.89-0.99)** |
| **Time from Onset** |  |  |  |  |  |  |  |  |  |  |  |  |
| ≤12 weeks | **48.4**<br>**(7.8)** | **51.3**<br>**(7.5)** | **1.06**<br>**(1.02-1.10)** | **50.0**<br>**(7.8)** | **52.1**<br>**(6.9)** | **1.04**<br>**(1.00-1.08)** | **48.6**<br>**(11.1)** | **44.7**<br>**(9.2)** | **0.92**<br>**(0.87-0.97)** | **43.0**<br>**(8.8)** | **40.2**<br>**(7.1)** | **0.93**<br>**(0.89-0.98)** |
| >12 weeks | 53.3<br>(6.6) | 52.6<br>(6.7) | 0.99<br>(0.96-1.01) | 53.3<br>(6.4) | 52.2<br>(7.6) | 0.98<br>(0.95-1.01) | 46.0<br>(9.6) | 44.5<br>(9.8) | 0.97<br>(0.92-1.01) | 39.3<br>(7.0) | 39.8<br>(7.2) | 1.01<br>(0.98-1.05) |
| **Body Mass Index** |  |  |  |  |  |  |  |  |  |  |  |  |
| Normal | 55.1<br>(6.9) | 55.4<br>(7.0) | 1.01<br>(0.96-1.06) | 54.4<br>(5.5) | 54.7<br>(5.3) | 1.00<br>(0.97-1.05) | 45.1<br>(11.4) | 42.8<br>(9.0) | 0.95<br>(0.87-1.03) | 37.4<br>(5.8) | 37.4<br>(5.2) | 1.00<br>(0.95-1.05) |
| Overweight | 53.4<br>(5.7) | 53.9<br>(6.0) | 1.01<br>(0.97-1.04) | 54.2<br>(5.6) | 54.3<br>(6.0) | 1.00<br>(0.96-1.04) | 45.4<br>(9.3) | 42.6<br>(9.5) | 0.94<br>(0.88-1.00) | 38.2<br>(5.9) | 38.2<br>(5.5) | 1.00<br>(0.96-1.04) |
| Obese | 48.4<br>(7.5) | 49.4<br>(7.0) | 1.02<br>(0.98-1.06) | 49.5<br>(7.9) | 50.2<br>(7.8) | 1.01<br>(0.98-1.05) | **49.2**<br>**(9.4)** | **46.5**<br>**(9.5)** | **0.94**<br>**(0.90-0.99)** | **44.1**<br>**(8.8)** | **42.0**<br>**(7.8)** | **0.95**<br>**(0.91-1.00)** |
| **Education** |  |  |  |  |  |  |  |  |  |  |  |  |
| High school or less | 49.6<br>(6.4) | 48.9<br>(7.8) | 0.99<br>(0.92-1.06) | 50.6<br>(7.8) | 49.5<br>(8.5) | 0.98<br>(0.90-1.07) | 47.3<br>(11.9) | 45.4<br>(10.2) | 0.96<br>(0.86-1.07) | 41.9<br>(7.4) | 42.4<br>(8.6) | 1.01<br>(0.92-1.11) |
| Some college | **48.1**<br>**(8.0)** | **50.7**<br>**(6.9)** | **1.05**<br>**(1.01-1.10)** | **49.4**<br>**(8.3)** | **51.8**<br>**(7.4)** | **1.05**<br>**(1.00-1.10)** | 49.2<br>(11.7) | 46.3<br>(9.8) | 0.94<br>(0.88-1.01) | **44.0**<br>**(9.2)** | **40.8**<br>**(7.4)** | **0.93**<br>**(0.88-0.98)** |
| Bachelor or Advanced Degree | 53.4<br>(6.7) | 53.6<br>(6.7) | 1.00<br>(0.98-1.03) | 53.6<br>(6.0) | 53.3<br>(6.4) | 0.99<br>(0.97-1.02) | **45.9**<br>**(9.1)** | **43.2**<br>**(8.8)** | **0.94**<br>**(0.90-0.98)** | 38.9<br>(6.8) | 38.7<br>(6.1) | 0.99<br>(0.96-1.03) |
| **Flu Vaccine** |  |  |  |  |  |  |  |  |  |  |  |  |
| Unvaccinated | **49.5**<br>**(7.8)** | **52.0**<br>**(6.4)** | **1.05**<br>**(1.01-1.09)** | **50.7**<br>**(7.9)** | **53.1**<br>**(6.5)** | **1.05**<br>**(1.00-1.09)** | **48.9**<br>**(10.3)** | **44.6**<br>**(9.4)** | **0.91**<br>**(0.86-0.97)** | 41.7<br>(9.2) | 39.5<br>(6.7) | 0.95<br>(0.90-1.00) |
| Vaccinated | 52.6<br>(7.0) | 51.8<br>(7.6) | 0.98<br>(0.96-1.01) | 52.9<br>(6.6) | 51.6<br>(7.6) | 0.98<br>(0.95-1.01) | 46.0<br>(10.1) | 44.6<br>(9.5) | 0.97<br>(0.93-1.01) | 40.0<br>(7.2) | 40.3<br>(7.3) | 1.01<br>(0.97-1.04) |
| **Smoking** |  |  |  |  |  |  |  |  |  |  |  |  |
| Non-Smoker | 52.1<br>(7.2) | 52.0<br>(7.2) | 1.00<br>(0.97-1.02) | 52.6<br>(6.7) | 52.1<br>(7.3) | 0.99<br>(0.97-1.02) | **46.4**<br>**(9.8)** | **44.4**<br>**(9.4)** | **0.96**<br>**(0.92-0.99)** | 40.1<br>(7.4) | 39.9<br>(7.1) | 1.00<br>(0.97-1.03) |
| Somedays/ Everyday | 47.0<br>(7.9) | 48.5<br>(5.8) | 1.03<br>(0.93-1.15) | 48.1<br>(9.3) | 53.1<br>(6.6) | 1.11<br>(0.98-1.24) | 51.7<br>(13.1) | 48.0<br>(10.4) | 0.93<br>(0.79-1.09) | 45.5<br>(10.3) | 42.4<br>(8.2) | 0.93<br>(0.81-1.07) |
| **Underlying Conditions** |  |  |  |  |  |  |  |  |  |  |  |  |
| None | 53.8<br>(6.3) | 53.7<br>(6.7) | 1.00<br>(0.97-1.03) | 53.9<br>(5.3) | 53.5<br>(6.1) | 0.99<br>(0.96-1.02) | 45.5<br>(10.4) | 43.4<br>(8.4) | 0.95<br>(0.90-1.00) | 38.5<br>(6.2) | 38.8<br>(6.3) | 1.01<br>(0.97-1.05) |
| 1 or more | 50.0<br>(7.7) | 50.6<br>(7.2) | 1.01<br>(0.98-1.04) | 50.9<br>(7.9) | 51.2<br>(7.8) | 1.01<br>(0.98-1.04) | **48.0**<br>**(9.9)** | **45.4**<br>**(10.0)** | **0.95**<br>**(0.91-0.99)** | 42.1<br>(8.6) | 40.8<br>(7.5) | 0.97<br>(0.93-1.01) |
| **Self-reported general health status** |  |  |  |  |  |  |  |  |  |  |  |  |
| Excellent | 57.1<br>(6.0) | 57.1<br>(6.6) | 1.00<br>(0.96-1.04) | 54.9<br>(5.0) | 55.3<br>(4.4) | 1.01<br>(0.98-1.04) | 44.2<br>(10.4) | 41.8<br>(7.8) | 0.95<br>(0.88-1.01) | 36.7<br>(4.7) | 36.8<br>(4.7) | 1.01<br>(0.97-1.04) |
| Very good or Good | 50.7<br>(6.3) | 50.5<br>(6.4) | 1.00<br>(0.97-1.02) | 51.9<br>(6.9) | 51.4<br>(7.4) | 0.99<br>(0.96-1.02) | 47.0<br>(9.4) | 45.1<br>(9.5) | 0.96<br>(0.92-1.00) | 41.2<br>(7.8) | 41.0<br>(7.4) | 1.00<br>(0.96-1.03) |
| Fair or Poor | 42.2<br>(7.5) | 41.3<br>(7.2) | 0.98<br>(0.82-1.17) | 46.0<br>(10.0) | 43.7<br>(12.6) | 0.95<br>(0.76-1.19) | 55.0<br>(12.7) | 57.4<br>(12.9) | 1.04<br>(0.84-1.29) | 47.9<br>(10.0) | 45.1<br>(7.0) | 0.94<br>(0.77-1.15) |

CI, confidence interval; SD, standard deviation
<sup>1</sup> Higher T-score represents more of the concept being measured. For negatively worded questions, a T-score of 60 is one standard deviation worse than average, and T-score of 40 is one standard deviation better than average.
<sup>2</sup> Calculated as mean T-score for participants in the SARS-CoV-2 test-positive group divided by mean T-score among participants in the SARS-CoV-2 test-negative group. Statistically significant mean T-score ratios are highlighted in bold text.
<sup>3</sup> Higher score corresponds to better health
<sup>4</sup> Higher score corresponds to greater limitation or more fatigue.

**Supplemental Table 3.** Multivariable Gamma regression mean T-score ratios and 95% confidence intervals from subgroup and sensitivity analyses. Models adjusted for base factors and health factors.

|  | **Positive SARS-CoV-2 test result (N)** | **Negative SARS-CoV-2 test result (N)** | **Global Health** | **Physical Function** | **Fatigue** | **Dyspnea** |
| --- | --- | --- | --- | --- | --- | --- |
| Primary analysis | 312 | 266 | 1.01 (0.99, 1.03) | 1.01 (0.98, 1.03) | 0.96 (0.93, 1.00) | 0.99 (0.96, 1.01) |
| Participants who reported fever at enrollment | 208 | 132 | 0.99 (0.96, 1.01) | 1.00 (0.97, 1.03) | 0.98 (0.93, 1.03) | 0.99 (0.95, 1.02) |
| Participants with $\geq 3$ underlying health conditions | 62 | 61 | 1.00 (0.95, 1.06) | 0.99 (0.93, 1.06) | 1.01 (0.94, 1.09) | 0.97 (0.90, 1.04) |
| Seropositive participants<sup>1</sup> excluded | 138 | 46 | 1.02 (0.98, 1.06) | 1.04 (0.99, 1.09) | 0.97 (0.90, 1.05) | 0.95 (0.89, 1.00) |
| Participants who reported having recovered at first follow-up<sup>2</sup> excluded | 115 | 69 | 0.99 (0.96, 1.03) | 1.00 (0.95, 1.04) | 0.96 (0.90, 1.03) | 0.99 (0.94, 1.05) |

<sup>1</sup> Participants at the Wisconsin site who tested RT-PCR negative for SARS-CoV-2 at enrollment and had serologic evidence of SARS-CoV-2 infection 28–42 days after illness onset
<sup>2</sup> Participants who responded “yes” to the question “Have you fully or mostly recovered from your illness?” on the first follow-up survey 7–14 days after illness onset.

